# Monitoring and evaluation of community interventions for viral hepatitis among migrants and refugees: a Delphi-based study

**DOI:** 10.1101/2025.05.08.25326721

**Authors:** Domenico Pascucci, Aina Nicolas, Abdelrahman Taha, Jeffrey V Lazarus, Matteo Di Pumpo, Vittoria Tricomi, Francesco Di Berardino, Carlo La Vecchia, José A Perez-Molina, Giuseppe Colucci, Camila A Picchio, Angelo Maria Pezzullo, Stefania Boccia

**Affiliations:** Section of Hygiene, Department of Life Sciences and Public Health, Università Cattolica del Sacro Cuore, Rome, Italy; Fondazione Policlinico Universitario A. Gemelli IRCCS, Rome, Italy; Barcelona Institute for Global Health (ISGlobal), Barcelona, Spain; CUNY Graduate School of Public Health and Health Policy, New York, NY, USA; Department of Clinical Sciences and Community Health, University of Milan, Milan, Italy; National Referral Centre for Tropical Diseases, Infectious Diseases Department, Hospital Universitario Ramón y Cajal IRYCIS, Madrid, Spain; CIBER de Enfermedades Infecciosas, Instituto de Salud Carlos III, Madrid, Spain; Division of Gastroenterology and Hepatology, Foundation IRCCS Ca’ Granda Ospedale Maggiore Policlinico, Milan, Italy

**Author notes:** co-first authors (Domenico Pascucci, Aina Nicolas). co-senior authors (Camila A Picchio, Angelo Maria Pezzullo, Stefania Boccia). Correspondence to: Angelo Maria Pezzullo, MD, MSc, PhD.

**Keywords:** infectious disease control, marginalised populations, viral hepatitis, point□of□care testing, linkage to care, vaccination coverage, morbidity, health system metrics

## Abstract

**Introduction:** Migrants and refugees in Europe carry a disproportionate burden of chronic hepatitis B and C and face barriers accessing formal health systems. Community-based interventions can improve screening, prevention, and care, yet no framework exists to track the performance of these programmes. This study aimed to generate a consensus set of indicators for monitoring and evaluating such interventions.

**Methods:** A scoping review (PubMed, January 2005□June 2024) identified 70 studies and 275 candidate indicators. After removing redundancies, we submitted 38 primary and 17 additional indicators to a two□round online Delphi panel. Fourteen experts in viral hepatitis, migrant health and community programmes rated each indicator on relevance, measurability, accuracy, ethics and clarity (4□point Likert scale). Items with >67 % combined “agree/somewhat agree” advanced. In Round 2, revised indicators were re□rated and all accepted indicators domain□ranked. Agreement grades (U, A, B, C) reflected the final proportion of positive votes; mean ranks identified domain priorities.

**Results:** All 38 primary indicators and 10 of 17 additional indicators surpassed the Round 1 threshold. After expert feedback, 15 indicators were re□rated in Round 2; none were rejected. The final set comprised 50 indicators distributed across six domains: Prevention (6), Testing (9), Linkage to care (6), Treatment & Care (9), Morbidity (7) and Health System (13). Overall combined agreement averaged 95.3% (SD 7.0), with 29 indicators achieving unanimous support. Testing and Morbidity domains showed the strongest consensus. Ranking highlighted screening acceptability, infection prevalence, rapid testing results, referral success and treatment initiation as highest priorities. An 18□indicator core subset (top three per domain) was proposed for routine use.

**Conclusion:** This Delphi study delivers the first consensus□driven indicator suite for monitoring and evaluating community hepatitis B/C services targeting migrants and refugees. Adoption of the 50□indicator framework, and its streamlined core set, can harmonise monitoring, guide resource allocation and strengthen data□driven progress toward elimination goals.

**KEY MESSAGES:** *What is already known on this topic:* Community□based HBV/HCV initiatives can reach migrants and refugees who face barriers to facility□based care, yet there is no agreed set of indicators to gauge the performance of these programmes, hindering comparability and quality improvement.

*What this study adds:* Using a two□round Delphi process, experts reached high consensus on 50 monitoring indicators (of which 18□indicator form a core subset) covering prevention, testing, linkage, treatment, morbidity and health□system support specifically for migrant□ and refugee□focused community services.

*How this study might affect research, practice or policy:* Adopting these indicators can standardise evaluation, guide resource allocation and enable data□driven adjustments to accelerate progress toward viral hepatitis elimination targets among underserved mobile populations.

## INTRODUCTION

An estimated 354 million people live with chronic hepatitis B or C worldwide, potentially exposing them to cirrhosis, hepatocellular carcinoma (HCC) and premature death (1). To curb this burden, in 2016, the World Health Organization (WHO) set a 2030 elimination target, which includes a reduction in incident cases by 90% and a reduction in mortality by 65% (2), yet, without intensified action, 20 million additional deaths are expected between 2015 and 2030 (3).

Although diagnostic and therapeutic advances have transformed liver disease care during the past 30□years (4), over five□million European Union/European Economic Area (EU/EEA)residents still have chronic viral hepatitis conditions□ (5). Globally, HBV and HCV cause 57% of cirrhosis cases and, together with liver cancer, account for 3.5% of all deaths; cirrhosis alone ranks 11th among causes of mortality and third for people aged□45□64 (5,7). The highest endemicity remains in sub□Saharan Africa and parts of Asia, where HBV prevalence reaches 5□6□% and HCV about 2% (9)

Migration from these high□prevalence regions is reshaping Europe’s epidemiology (10). More than half of migrants originate from HBV□endemic areas and nearly 80□% from HCV□endemic countries. Among individuals arriving from intermediate□ or high□prevalence settings, HBsAg and anti□HCV positivity reach 6□% and 2.3□%, respectively, with local studies reporting HBV rates up to 32□% in Italy and 21□% in Spain□(10)(12). Migrants now represent roughly a quarter of chronic HBV and 14□% of chronic HCV cases in the EU/EEA—about 2□million and 0.9□million infections, respectively (11).

Responding to this challenge and echoing “Europe’s□Beating Cancer” plan (13), the EU4Health co–funded Multi-country Viral Hepatitis COMmunity Screening, Vaccination, and Care (VH-COMSAVAC) project operated in Greece, Italy and Spain, where migrant and refugee communities carry disproportionate HBV/HCV burdens (14–16). Its model combines point□of□care testing, decentralised HBV vaccination and streamlined referral to specialist services for those with reactive results, aiming to avert future morbidity and mortality driven by undiagnosed infection.

The COVID□19 pandemic strengthened the case for community□based health delivery. The WHO, the International Federation of Red Cross and Red Crescent Societies, and UNICEF emphasized how empowered communities accelerate uptake of new vaccines, diagnostics and treatments (17,18). For hepatitis, community-based strategies, extend reach beyond traditional facilities, detect infection earlier and serve groups chronically underserved by formal systems (9,19). By integrating rapid testing, on□site vaccination, culturally adapted education and peer□supported linkage to care, they mitigate barriers such as cost, waiting times, language, legal insecurity and stigma (9,20–25). Consequently, programmes targeting migrants or people who use drugs often report both high coverage and high positivity rates (22).

Continuous performance monitoring of community-based strategies is critical to identify gaps, replicate success and judge scalability. Existing indicator frameworks, such as the WHO strategic information cascade (26), and the European Centre for Disease Prevention and Control (ECDC) monitoring system (27), were not specifically designed for community settings. Tailored metrics are therefore needed to capture the full pathway from outreach and vaccination to diagnosis, linkage, treatment and cure in non□clinical contexts.

This article describes the Delphi□based process used within VH□COMSAVAC to define key performance indicators for monitoring and evaluation strategies for HBV/HCV screening, prevention, and management among migrants and refugees at the community level. The resulting set of indicators seeks to support data□driven evaluation, inform policy and advance Europe’s contribution to the 2030 hepatitis elimination goal.

## METHODS

This study used a Delphi consensus method to identify and to prioritize key performance indicators (KPIs) for monitoring and evaluating community-based viral hepatitis (HBV and HCV) screening, prevention, and management strategies. The process began with a scoping review of the literature through which an initial set of indicators which were identified and later refined to avoid redundancy. Experts then prioritized this list through Delphi rounds. The research protocol, including detailed methods, was registered on OSF (28); amendments and clarifications appear in Supplementary Materials.

### Scoping literature review

#### Search strategy

A scoping review identified indicators for monitoring and evaluating HBV and HCV screening, prevention, and management in community-based strategies among migrants and refugees; HIV programme indicators were also examined for transferable insights. Community-based interventions were defined as those providing decentralized healthcare services, such as communicable disease screening, outside formal healthcare settings. These efforts, incorporating preventive measures, aim to improve public health responses and complement facility-based care, which typically emphasizes on curative and rehabilitative services (29). The review followed the Arksey and O’Malley framework (30) and the Joanna Briggs Institute manual (31). PubMed (January 2005-June 2024) was searched with terms such as "HBV," "HCV," "indicat*," "screen*," "commun*," and "migra*". The complete search string can be found in the protocol (28) for further reference. Reporting complied with the Preferred Reporting Items for Systematic Reviews and Meta-Analyses extension for Scoping Reviews (PRISMA-ScR)(32).

#### Study selection

Two groups of researchers (DP and FDB; AN and VT) independently screened titles and abstracts; full-text evaluations were then divided among the four researchers, and disagreements were resolved by a third reviewer (CAP). Inclusion criteria required studies published in English from 2005 onwards, focusing on community-based interventions for HBV, HCV, or HIV, targeting migrants or refugees, and including at least one well-described performance indicator. Articles not meeting these criteria or with incomplete data were excluded. Rayyan software (33) was used for screening.

#### Data extraction

Two reviewers (DP and AN) independently extracted the data using an Excel sheet, recording bibliographic details and country, disease focus, target population, indicator title with a brief description if available. All indicators that were explicitly described in the selected studies, as well as those derived from available reported data, were extracted. Tables, figures, and supplementary materials were also examined.

#### Selection of initial performance indicators

Indicators that were semantically similar or calculated using analogous methods were eliminated to avoid redundancy. Indicators that were irrelevant to the community setting were also discarded. All decisions were made through consensus among two independent reviewers (DP, AN) to ensure clarity and precision in the initial set of indicators to be submitted to the Delphi process. When unsure about their inclusion, indicators were set aside to compile an additional list, and later, during the Delphi process, experts were consulted about their inclusion.

### Delphi consensus

At the conclusion of the selection process, each indicator was categorized into one of the following health domains: *prevention, testing, treatment and care, morbidity*, and *health system*. These domains were selected among those used by WHO in its technical guidelines for monitoring and evaluating HBV and HCV (26), based on their relevance to evaluating community-based interventions.

#### Prioritization of indicators

The Delphi method was chosen to prioritize the preliminary set of indicators identified by the core research team after reviewing the literature. This structured consensus technique involves iterative rounds of expert evaluation (34–36). A modified Delphi approach was applied, replacing the exploratory first round with a scoping review to generate an initial list of indicators for evaluation (37).

Experts with diverse backgrounds, including specialists in viral hepatitis and professionals with expertise in community-based programmes, migrant health, and public health, particularly in Europe, were recruited through a convenience sampling approach. Invitations outline main project objectives, evaluation criteria and time commitment; participation was voluntary, anonymous, and confidential, with 15-day response windows (extended by 16 and one day in the two rounds, respectively, exact dates in Results). Only complete responses were analysed.

#### Delphi method data collection

The study followed two rounds (Round 1 – R1 and Round 2 – R2) of consultation which were conducted using the online platform SurveyMonkey. In R1, panellists were asked to rate the indicators based on the following criteria (based on a similar prioritisation exercise (38,39)): relevance, measurability, accuracy, ethical considerations, and comprehensibility (Box 1), using a four-point Likert scale (“agree”, “somewhat agree”, “somewhat disagree”, and “disagree”) with an additional option for "not qualified to respond" (40–42). Sociodemographic and professional characteristics of panellists (e.g., gender, age, country of employment, job category and level of expertise) were systematically collected from participants in R1. Panellists also provided open-ended feedback to support the refinement of indicators, which was subsequently presented in aggregated form, without differentiation by specific professional group or discipline.

##### Box 1. Rating criteria for the indicators’ prioritisation exercise

RELEVANT - In a community setting, a suitable indicator must have a robust clinical and/or empirical basis justifying its use. It should provide valuable insights relevant to various stakeholders engaged in community-based practice and policy, thereby fostering efficient actions within this context.

MEASURABLE - Within a community framework, the data necessary for evaluating the indicator should be easily obtainable. This ensures that community-based programs can regularly assess and monitor their progress.

ACCURATE - A proper indicator in a community setting should reveal significant variations in the delivery of care (sub)-processes across different community services and/or regions. These differences must be meaningful and not merely the result of random fluctuations or characteristics of the community members.

ETHICAL - In a community context, the collection, management, and analysis of indicator data must uphold the individual rights to confidentiality and informed consent, and respect the freedom of choice regarding data provision. This is particularly important in community settings where there’s a close interaction with individuals and their personal data.

COMPREHENSIVE - An indicator in a community setting should be straightforward, ensuring that its interpretation is easily comprehensible not only to experts and stakeholders but also to the general community population. This clarity is crucial for effective communication and engagement in community-based initiatives

Indicators that achieved less than 67% (41,43) agreement for responses of "agree" or "somewhat agree" (combined agreement) in R1 were excluded from further consideration. Conversely, those exceeding this threshold proceeded to R2. Open-ended feedback from experts was revised and evaluated by the research team and used for the refinement of indicators before R2. Indicators that did not undergo substantial modifications advanced directly to the ranking phase, while those that underwent significant revisions were re-evaluated through an additional rating process during R2.

During R1, alongside the rating of the preliminary list of indicators, panellists were also asked to evaluate the additional list of indicators, comprising those for which uncertainty about inclusion emerged in the previous phase. Each of these indicators was assessed using a simple yes/no format to determine whether it should be included in the main list. Indicators that received at least 67% affirmative votes from the panel were added to the main list and subsequently rated in R2. Only R1 completers were invited to R2, ensuring consistency in the consensus process.

In R2, panellists reviewed the revised indicators along with summaries outlining the modifications made. Indicators that had undergone substantial changes following R1 feedback, as well as those selected from the additional list, were subjected to re-evaluation (re-rating). During the re-evaluation processes, using the same four-point Likert scale as in R1, panellists reassessed these indicators to determine their suitability for inclusion in the final list.

Separately, panellists were asked to rank the indicators within each health domain based on their practical applicability in monitoring and evaluation of community-based programs for viral hepatitis B and C screening, prevention, and management among migrants and refugees. For the ranking process, panellists arranged the indicators in order of relevance, from the most to the least important in their opinion. If an indicator was considered not relevant, they could choose the option “I prefer not to rank this indicator”. Experts were given the opportunity to suggest minor edits that enhanced the clarity of the indicator without changing its meaning, while feedback requiring substantial modifications was not considered. A systematic step-by-step list for the Delphi process is provided in Supplementary Materials.

#### Delphi data analysis

The quantitative analysis of R1 and R2 focused on the evaluation of each indicator using a 4-point Likert scale, applied across five predefined items. Each panellist assigned scores from 1 (“Disagree”) to 4 (“Agree”), with the option to select “Not qualified to respond.” For each indicator, the five item scores were summed to calculate an individual total score per panellist, ranging from 5 to 20. To classify the level of agreement, the total score was mapped onto one of the four predefined categories by dividing the score range (5–20) into four equal intervals of 3.75 points. When panellists selected “Not qualified to respond” for one or more items, those responses were excluded from the calculation, and the score range was proportionally adjusted based on the number of valid responses. Finally, the distribution of individual scores across the four categories was calculated for each indicator to assess the overall level of agreement.

After the completion of R2, a grading system was applied to each indicator to reflect the overall level of combined agreement (“Agree” and “Somewhat agree”) reached by the panel. This grading approach follows the convention of previous Delphi studies (34–36), with ‘U’ indicating unanimous agreement (100%), ‘A’ for 90–99%, ‘B’ for 78–89%, and ‘C’ for 67– 77%.(40–42).

For the ranking process, the mean score was used to summarise the scores assigned to each indicator. Since position 1 in the ranking represented the highest level of relevance, the ranking was structured so that indicators with the lowest mean scores—indicating greater perceived importance—were assigned higher positions. All responses were included in the calculation, including those where participants selected “I prefer not to rank this indicator.” In such cases, the lowest possible ranking value was assigned to those responses, so that indicators with a higher number of “Not to Rank” selections were penalised accordingly. Moreover, given the expected high number of indicators and the complexity of evaluating community-based interventions, the potential need to identify a more focused subset of KPIs to support practical application was acknowledged (44). As part of the methodological design, it was planned to use the results of the ranking process to guide the selection of the top three core indicators within each of the six predefined domains. While not exhaustive, this subset would aim to capture key aspects of each domain and serve as a proxy for the overall effectiveness of the intervention. No subgroup-specific analyses (e.g., based on professional background or discipline) or weighting of responses were applied during the data analysis. All experts’ ratings and rankings were considered equally in the calculation of agreement levels and final indicator prioritization.

The DELPHISTAR guideline was used for reporting and the compiled checklist is in Supplementary Materials (45). No external consultation occurred regarding the methodology. Descriptive analysis of panellist characteristics is in Supplementary Materials.

## RESULTS

### Scoping review and list of potential performance indicators

The literature search identified 2,354 studies and a total of 157 articles underwent full-text review, of these, 87 articles were excluded. The remaining 70 studies were included in the analysis. The screening process is detailed in the PRISMA flow diagram (Figure 1).

**Figure 1:**
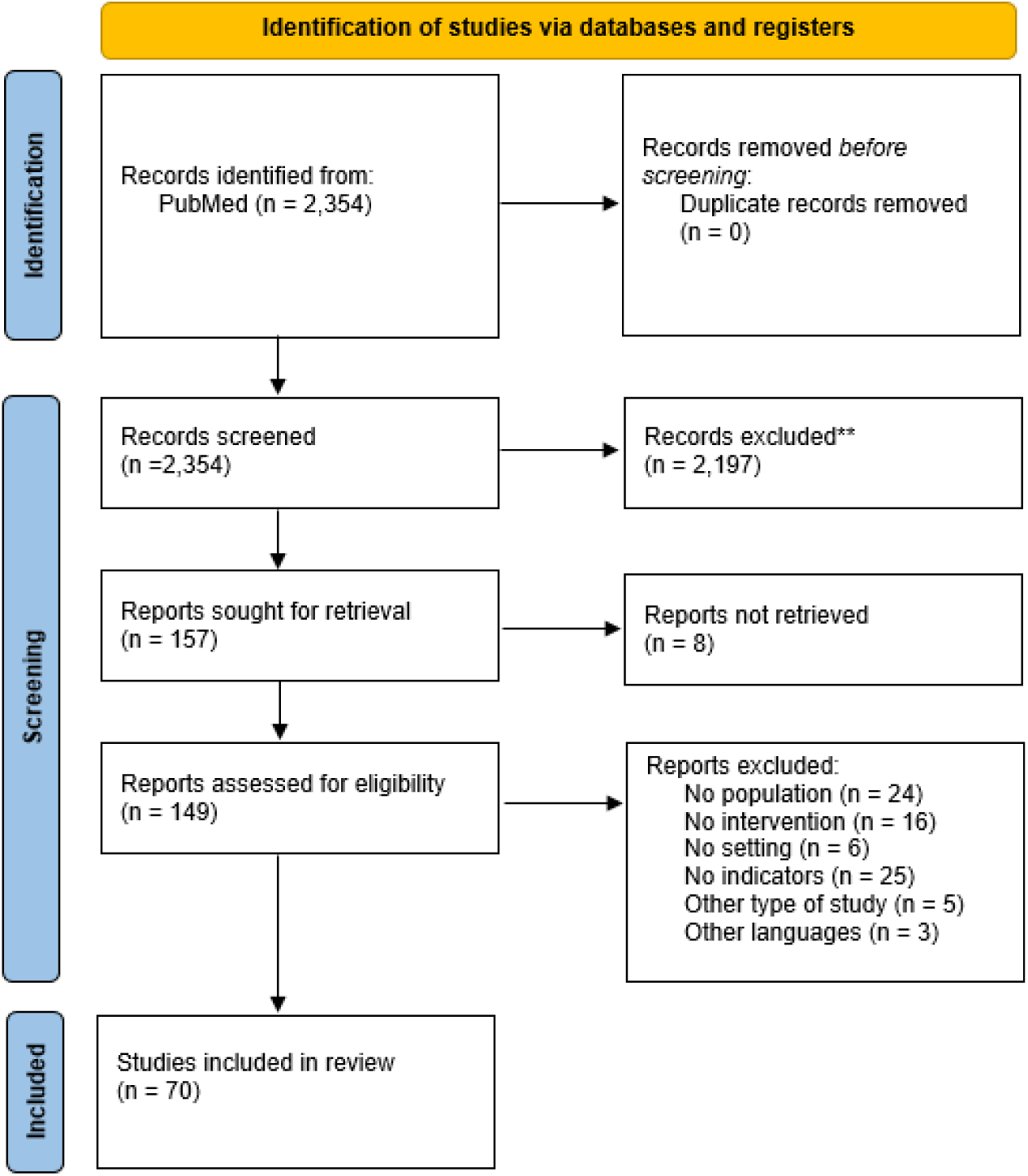
PRISMA flow diagram for the scoping review

A total of 275 indicators were extracted. The list of included studies derived from the scoping review and the initial list of 275 extracted indicators are available in Supplementary Materials. Redundant and irrelevant indicators were removed and a preliminary main list of 38 indicators plus an additional list of 17 indicators were included for expert evaluation (Supplementary Materials).

### Results of the Delphi consensus study

Following the selection process, the preliminary set of indicators was defined and grouped into six key health domains: *prevention, testing, linkage to care, treatment and care, morbidity*, and *health system*, as shown in Table 1.

**Table 1:** Distribution of Indicators across Health Domains in the Main and Additional Lists.

| Domains | Main list | Additional list |
| --- | --- | --- |
| *Prevention* | 6 indicators | 4 indicators |
| **Testing** | 6 indicators | 3 indicators |
| **Morbidity** | 5 indicators | 1 indicator |
| **Linkage to care** | 8 indicators | 1 indicator |
| **Treatment & Care** | 7 indicators | 3 indicators |
| **Health System** | 6 indicators | 5 indicators |

#### Round 1

R1 took place from December 6, 2024, to January 6, 2025, following an email sent to panellists providing detailed instructions. A total of 41 experts, previously identified by the research group, were invited to participate. The response rate was 85% (35 out of 41 experts), however, only the 14 experts (34%) provided complete responses and were therefore considered in the analysis.

The expert panel comprised 14 participants (median age 38 years, inter-quartile range 30–45). Most were women (10/14; 71%). Panel members were based in six countries: Spain (5; 36%), the United Kingdom (3; 21%), Italy (2; 14%), Greece (2; 14%), and one each from Belgium and the United States (both 7%). Almost two-thirds were employed in academia (9; 65%), with the remainder working in the public sector (3; 21%) or civil-society organisations (2; 14%). Their principal fields of work were public health (6; 43%), clinical research (5; 36%) and clinical practice (3; 21%). Regarding professional experience, four participants (29%) had 1–5 years in the field, three (21%) had 6–10 years, four (29%) had 11–15 years, and three (21%) had more than 20 years; none had 16–20 years of experience.

All 38 indicators in the preliminary main list received 67% or more agreement in responses marked as ‘agree’ or ‘somewhat agree’ (A+SA) in R1, with an overall average agreement (A+SA) of 93.8% (SD ± 8.2%). Among the domains, Morbidity achieved the highest agreement with a mean of 100.0% (SD ± 0.0%), while the Health System domain recorded the lowest with a mean of 84.5% (SD ± 7.6%). A detailed description of the results from R1 is available in Supplementary Materials.

Twenty indicators reached the highest level of agreement (100%). Examples for these unanimously accepted indicators are: *“Screening acceptability”*, *“HBV DNA testing”* (among all HBsAg positives), *“Prevalence of active HCV infection”*, and proportions of participants receiving communication of rapid- and laboratory tests’ results. The indicators with the lowest proportion of agreement were “*Cost per test-positive participant”*, “*Risk behaviour”, “Sustained Virologic Response at 12 weeks/24 weeks”* and “*Sampling rate”*, all with 78.6% agreement.

Regarding the indicators in the additional list, 10 indicators received at least 67% affirmative votes and were added to the main list, subsequently rated during R2.

#### Analysis of expert comments and feedback

Following the analysis of open-ended feedback from Round 1, we reviewed and revised several indicators. Among the indicators in the main preliminary list, only one did not receive any qualitative comments. Of the remaining indicators, eight were suggested for reallocation to a different domain. Minor modifications were applied to 27 indicators, including modifications in their names or definitions, which did not imply a modification in their meaning nor their calculation methods. Additionally, two indicators, “*HBsAg-Positive participants with detectable HBV-DNA”* and “*Prevalence of active HCV infection”*, were reassigned from the *Testing* to the *Morbidity* domain to better reflect their role in measuring disease burden. Given the non-substantial nature of these changes, none of these indicators were re-evaluated through a rating process in R2.

Five indicators underwent substantial modifications and were re-evaluated in R2. Among them, “*Acceptability rate (counselling and screening)”* was renamed “*Education uptake”* to better reflect its focus on participation in education programs. Experts noted that counselling was not the most appropriate term and that screening acceptability was already assessed separately.

To clarify the role of DNA/RNA testing in community settings, two new indicators were introduced, as per the feedback from three experts: “*HBV DNA testing”* and “*HCV RNA testing”*, which measure the proportion of eligible participants offered these tests. These were added to the *Testing* domain following the reclassification of “*HBsAg-Positive participants with detectable HBV-DNA”* and “*Prevalence of active HCV infection”* into the *Morbidity* domain.

In the *Health System* domain, “*Cultural mediators - Translators - Peer navigators”* was split into “*Ratio of participants per cultural mediator, translator and/or peer navigator”*, which quantifies the number of mediators and translators involved, and “*Participant use of mediation services”*, which measures participant uptake of these services. This distinction clarifies the difference between personnel availability and service utilization. The synthesis of the results of the post-R1 analysis are reported in Table 2. A total of 50 indicators progressed to R2, with 15 requiring re-rating due to substantial changes or being assessed for the first time, while 35 indicators proceeded directly to the ranking phase. The list of indicators presented in R2 is available in Supplementary Materials.

**Table 2:** Results of the Analysis of Expert Comments and Feedback after Round 1.

| **Domains** | **Indicators confirmed at first round (no substantial change)** | **Indicators requiring re-rating due to substantial change** | **First-time rating for indicators from additional list** | **Total number of indicators** |
| --- | --- | --- | --- | --- |
| **Prevention** | 5 | 1 | 3 | 9 |
| **Testing** | 4 | 2 | 3 | 9 |
| **Morbidity** | 7 | 0 | 0 | 7 |
| **Linkage to care** | 7 | 0 | 1 | 8 |
| **Treatment & Care** | 7 | 0 | 1 | 8 |
| **Health System** | 5 | 2 | 2 | 9 |
| **Total** | 35 | 5 | 10 | 50 |

#### Round 2

R2 was conducted between February 7 and February 23, 2025, and all 14 participants from R1 completed R2. All 15 indicators subjected to re-rating surpassed the supermajority threshold of A+SA, achieving a mean proportion of agreement of 97.2% (SD ± 3.5%).

A detailed description of the results obtained in R2 is available in Supplementary Materials.

Following R2, 12 indicators which underwent re-rating obtained open-ended feedback which required revision by experts to consider potential edits. Minor edits that enhance the clarity of the indicator were made based on expert feedback for the following indicators: “*Education uptake”*, “*HBV immunity prevalence”*, “*Additional assessment among positive participants”* (*Linkage to care*), “*Additional assessment among positive participants”* (*Treatment and Care*), and “*Ratio of participants per cultural mediator, translator, and/or peer navigator”*.

Table 3 presents the final list of 50 indicators, their grade of agreement, and the rankings for each of the six domains.

**Table 3:** Final list of indicators with grade of agreement and ranking.

| FINAL LIST |  |  |  |  |  |  |  |  |  |
| --- | --- | --- | --- | --- | --- | --- | --- | --- | --- |
|  | Indicator | Definition | GRADE | A (%) | SA (%) | SD (%) | D (%) | NQ (%) | RANKING |
| **PREVENTION** |  |  |  |  |  |  |  |  |  |
| 1 | **Education uptake** | Proportion of participants who attended educational* session/s (as part of the programme) among all participants<br>*\*Education session/s focused on the prevention, early screening, diagnosis, treatment, and care of viral hepatitis infections* | **A** | 71.5 | 21.4 | 0 | 0 | 7.1 | **4** |
| 2 | **Screening acceptability** | Proportion of participants who agreed to be screened for HBV/HCV among participants who were offered screening | **U** | 92.9 | 7.1 | 0 | 0 | 0 | **1** |
| 3 | **Self-reported previous testing** | Proportion of participants who self-reported having been previously screened for HBV and/or HCV among all screened participants | **B** | 50.0 | 35.7 | 14.3 | 0 | 0 | **9** |
| 4 | **Self-reported Risk Behaviours** | Proportion of participants who self-report having engaged in risk behaviours* among all screened participants<br>*\* Risk behaviour refer to activities or practices which increase the likelihood of acquiring HBV and/or HCV, including: unprotected sex, sharing of needles, use of non-sterilized material for e.g., tattoo, traditional scarring, or in medical practices* | **B** | 50.0 | 28.6 | 14.3 | 7.1 | 0 | **8** |
| 5 | **HBV vaccination coverage (at least one dose)** | Proportion of participants who received at least one dose of the HBV vaccine (as part of the programme) among those eligible for and offered HBV vaccination | **B** | 64.3 | 21.4 | 14.3 | 0 | 0 | **2** |
| 6 | **HBV vaccination coverage (complete regime)** | Proportion of participants who received all required HBV vaccine doses (as part of the programme) among those eligible for and offered HBV vaccination | **A** | 71.5 | 21.4 | 7.1 | 0 | 0 | **2** |
| 7 | **Acceptability of resources and education program** | Median score rating (0-5) for: (1) leaflet readability, leaflet relevance, (2) acceptability questionnaire at follow-up, and (3) clarity of in-person explanation, and (4) useful/relevant information of in-person explanation | **U** | 35.7 | 64.3 | 0 | 0 | 0 | **7** |
| 8 | **Changes in knowledge and understanding (self-reported and/or questionnaire)** | Proportion of participants who reported a better understanding or increased knowledge of HBV/HCV after educational session/s among all participants | **U** | 71.4 | 28.6 | 0 | 0 | 0 | **5** |
| 9 | **Self-reported previous HBV vaccination** | Proportion of participants reporting previous HBV vaccination among all screened participants | **A** | 64.3 | 28.6 | 7.1 | 0 | 0 | **6** |
| **TESTING** |  |  |  |  |  |  |  |  |  |
| 1 | **Prevalence of HBV infection** | Proportion of participants with a reactive HBsAg test among all screened participants | **U** | 85.7 | 14.3 | 0 | 0 | 0 | **1** |
| 2 | **Prevalence of current or past HCV infection (based on HCV-antibody testing)** | Proportion of participants with a reactive anti-HCV test among all screened participants | **U** | 85.7 | 14.3 | 0 | 0 | 0 | **2** |
| 3 | **Participants unaware of their HBV and/or HCV status** | Proportion of participants who were unaware of their status among those with a reactive HBV and/or HCV test (i.e., HBsAg+ and/or antiHCV+) | **U** | 71.4 | 28.6 | 0 | 0 | 0 | **7** |
| 4 | **HBV DNA testing** | Proportion of HBsAg positive participants who were offered HBV DNA testing among all HBsAg positive participants | **U** | 100.0 | 0 | 0 | 0 | 0 | **4** |
| 5 | **HCV RNA testing** | Proportion of anti-HCV positive participants who were offered HCV RNA testing among all anti-HCV positive participants | **U** | 92.9 | 7.1 | 0 | 0 | 0 | **3** |
| 6 | **Participant at risk of HBV infection** | Proportion of participants who tested negative for HBsAg and had no evidence of past HBV vaccination nor infection (self-reported or anti-HBs/anti-HBc negative) among all screened participants | **A** | 78.6 | 14.3 | 7.1 | 0 | 0 | **8** |
| 7 | **Serological evidence of previous HBV vaccination** | Proportion of participants positive for anti-HBs (and HBsAg- and anti-HBc-) among all screened participants | **U** | 100.0 | 0 | 0 | 0 | 0 | **6** |
| 8 | **Evidence of past resolved HBV infection** | Proportion of participants testing positive for anti-HBc (within HBsAg-negative) among all screened participants | **U** | 92.9 | 7.1 | 0 | 0 | 0 | **8** |
| 9 | **HBV immunity prevalence** | Proportion of HBsAg- participants who show evidence of immunity from a past resolved infection (anti-HBc+) or from previous vaccination (anti-HBs+) among all screened participants | **U** | 100.0 | 0 | 0 | 0 | 0 | **5** |
|  | **MORBIDITY** |  |  |  |  |  |  |  |  |
| 1 | **Prevalence of HBV-HCV Coinfection** | Proportion of participants who are simultaneously actively infected with HCV (anti-HCV+ and detectable HCV RNA) and HBV (i.e., HBsAg+) among all participants | **U** | 92.9 | 7.1 | 0 | 0 | 0 | **4** |
| 2 | **Prevalence of HBV-HDV Coinfection** | Proportion of participants with an active HBV infection (i.e., HBsAg+) and active HDV infection (anti-HDV+ and HDV RNA+) among all participants positive for HBsAg | **U** | 92.9 | 7.1 | 0 | 0 | 0 | **6** |
| 3 | **Prevalence of HCV-HIV Coinfection** | Proportion of participants who are simultaneously actively infected with HCV (anti-HCV+ and HCV RNA) and HIV among all participants with an active HCV infection (i.e., detectable HCV RNA) | **U** | 92.9 | 7.1 | 0 | 0 | 0 | **7** |
| 4 | **HBsAg-Positive participants with detectable HBV-DNA** | Proportion of HBsAg+ participants with detectable HBV- DNA among all participants testing positive for HBsAg | **A** | 71.5 | 21.4 | 7.1 | 0 | 0 | **5** |
| 5 | **Prevalence of active HCV infection** | Proportion of participants testing positive for anti-HCV and having detectable HCV-RNA among all screened participants | **U** | 100.0 | 0 | 0 | 0 | 0 | **1** |
| 6 | **Prevalence of end-stage liver disease** | Proportion of end-stage liver disease* cases detected among all participants positive for HBsAg<br>* *End-stage liver* *disease refers to chronic liver failure, which usually (but not necessarily) results from cirrhosis.* | **U** | 71.4 | 28.6 | 0 | 0 | 0 | **3** |
| 7 | **Prevalence of liver cancer** | Proportion of liver cancer (i.e., hepatocellular carcinoma - HCC) cases detected among all participants positive for HBsAg | **U** | 85.7 | 14.3 | 0 | 0 | 0 | **2** |
|  | **LINKAGE TO CARE** |  |  |  |  |  |  |  |  |
| 1 | **Onsite communication of rapid test results** | Proportion of participants receiving their screening test results (i.e., rapid test results) onsite among all screened participants | **U** | 92.9 | 7.1 | 0 | 0 | 0 | **1** |
| 2 | **Communication of results (laboratory-based tests)** | Proportion of participants receiving laboratory-based test* results in community setting among all screened participants<br>*\*tests requiring processing of blood samples outside immediate community setting* | **U** | 100.0 | 0 | 0 | 0 | 0 | **4** |
| 3 | **Referral of positive participants** | Proportion of participants with an active HBV infection (i.e., HBsAg+) and/or anti-HCV+ referred to specialist care among all HBsAg+and/or anti-HCV+ participants | **U** | 92.9 | 7.1 | 0 | 0 | 0 | **2** |
| 4 | **Linkage to care among positive participants** | Proportion of participants with an active HBV infection HBV infection (i.e., HBsAg+) or anti-HCV+ successfully linked to specialist care (with a documented first visit in specialist care) among all HBsAg+ and/or anti-HCV+ participants | **U** | 78.6 | 21.4 | 0 | 0 | 0 | **3** |
| 5 | **Linkage to care to collaborating centers** | Proportion of participants with an active HBV infection (i.e., HBsAg+) or anti-HCV+ successfully linked to specialist care at a clinical center linked to the project among all HBsAg+ and/or anti-HCV+ participants | **A** | 64.3 | 28.6 | 7.1 | 0 | 0 | **5** |
| 6 | **HBV vaccination offer** | Proportion of participants offered HBV vaccination among all participants eligible for HBV vaccination | **U** | 92.9 | 7.1 | 0 | 0 | 0 | **6** |
| 7 | **Linkage to care for post-test counselling for past resolved HBV infection** | Proportion of participants with a past resolved HBV infection receiving post-test counselling *among all participants with a past resolved HBV infection<br>*\* Post-test counselling refers to providing guidance to participants with resolved HBV infection, including health implications and follow-up recommendations.* | **U** | 71.4 | 28.6 | 0 | 0 | 0 | **8** |
| 8 | **Additional assessment among positive participants** | Proportion of HBsAg+ participants having at least one documented visit in specialist care and being assessed for ALT, HBV DNA levels, and liver cancer among all HBsAg+ participants | **A** | 71.5 | 21.4 | 0 | 7.1 | 0 | **7** |
| **TREATMENT AND CARE** |  |  |  |  |  |  |  |  |  |
| 1 | **HCV Treatment initiation** | Proportion of participants with an active HCV infection (anti-HCV+ and detectable HCV RNA) who started treatment among all participants with an active HCV infection | **U** | 92.9 | 7.1 | 0 | 0 | 0 | **1** |
| 2 | **HCV treatment completion** | Proportion of participants with an active HCV infection (anti-HCV+ and detectable HCV RNA) who completed HCV treatment among all participants with an active HCV infection who started HCV treatment | **U** | 92.9 | 7.1 | 0 | 0 | 0 | **2** |
| 3 | **HCV Treatment success** | Proportion of participants with an active HCV infection (anti-HCV+ and detectable HCV RNA) who finished HCV treatment and were cured among all participants with an active HCV infection who completed HCV treatment | **U** | 92.9 | 7.1 | 0 | 0 | 0 | **6** |
| 4 | **SVR12 - SVR24 following HCV treatment** | Proportion of participants with an active HCV infection (anti-HCV+ and detectable HCV RNA) who completed HCV treatment and achieved SVR at 12 and/or 24 weeks among all participants with an active HCV infection who completed HCV treatment | **B** | 78.6 | 0 | 0 | 0 | 21.4 | **5** |
| 5 | **HBV treatment eligibility** | Proportion of HBsAg+ participants who meet criteria* for initiating therapy among all HBsAg+ participants<br>*\* according guidelines corresponding to the region of implementation* | **A** | 71.5 | 21.4 | 7.1 | 0 | 0 | **4** |
| 6 | **HBV Treatment initiation** | Proportion of HBsAg+ participants initiating HBV treatment among all HBsAg+ participants who met the criteria for HBV treatment | **A** | 85.8 | 7.1 | 7.1 | 0 | 0 | **3** |
| 7 | **Retention in care** | Proportion of HBsAg+ participants with more than one appointment in specialist services between screening and end of follow-up period among all HBsAg+ participants | **U** | 57.1 | 42.9 | 0 | 0 | 0 | **6** |
| 8 | **Retention in care (6 months)** | Proportion of participants who had another visit (after the initial visit) 6 months after initial screening among all recruited participants | **A** | 92.9 | 0 | 7.1 | 0 | 0 | **8** |
| **HEALTH SYSTEM** |  |  |  |  |  |  |  |  |  |
| 1 | **Sampling proportion** | Proportion of recruited participants among the expected number of recruited participants | **B** | 42.9 | 35.7 | 21.4 | 0 | 0 | **2** |
| 2 | **Participation acceptability** | Proportion of participants recruited among all participants offered the opportunity to participate | **A** | 78.6 | 14.3 | 7.1 | 0 | 0 | **1** |
| 3 | **Self-reported barriers to access care** | Proportion of participants self-reporting encountering at least one barrier* in accessing care services among all recruited participants<br>*\*Barrier can be chosen (but are not limited to): language, cultural, financial, legal, structural and disinformation-related barriers.* | **A** | 78.6 | 7.1 | 14.3 | 0 | 0 | **3** |
| 4 | **Ratio of participants per cultural mediator, translator and/or peer navigator** | Ratio of participants per cultural mediator, translators and/or peer navigators involved in an intervention | **A** | 64.3 | 28.6 | 7.1 | 0 | 0 | **4** |
| 5 | **Participant use of mediation services** | Proportion of participants who utilized mediation services* provided by cultural mediators, translators and/or peer navigators among the total number of participants<br>*\*Mediation services may include translation, cultural adaptation services etc.* | **U** | 78.6 | 21.4 | 0 | 0 | 0 | **5** |
| 6 | **Intervention cost (per person)** | The total cost of the project (or stratified by type of activity) divided by the total number of recruited participants | **A** | 64.3 | 28.6 | 0 | 0 | 7.1 | **6** |
| 7 | **Cost per test-positive participant** | The total cost of the project (or stratified by type of activity) divided by the number of positive participants | **C** | 42.9 | 28.6 | 21.4 | 0 | 7.1 | **7** |
| 8 | **Recruitment capacity** | Number of sites visited for recruitment; Types of recruitment venues; Number visits for recruitment; Time spent in for recruitment. | **U** | 71.4 | 28.6 | 0 | 0 | 0 | **7** |
| 9 | **Screening and vaccination cost (per Person)** | Total cost of screening and vaccination activities divided by the total number of screened and vaccinated participants | **A** | 92.9 | 0 | 0 | 0 | 7.1 | **7** |

Grades are based on the percentage of combined agreement ('agree' + 'somewhat agree'). U, unanimous (100%) agreement; A, 90–99% agreement. Responses are presented as percentages of the total responses. A, agree; SA, somewhat agree; SD, somewhat disagree; D, disagree; NQ, the percentage of participants that indicated that they were not qualified to respond. Ranking scale: 1 represents the best evaluation, with higher numbers indicating progressively worse evaluations

Overall, the expert panel reached unanimous (“U”) combined agreement (“agree” + “somewhat agree”) for 29 indicators (58%), 15 indicators had a combined agreement between 90 and 99% (“A”), and 6 indicators (12%) had a combined agreement threshold below 90%. The mean level of combined agreement across all domains was 95.3% (SD ± 7.0%).

The *Testing* and *Morbidity* domains received the highest proportion of indicators classified as ‘U’ (unanimous agreement, 100%), with 88.9% and 85.7%, respectively. In contrast, *Prevention* and *Health System* had the lowest proportions of indicators assigned a ‘U’ grade, with 33.3% and 22.2%, respectively. Five indicators received a combined agreement corresponding to a “B” grade. One indicator received a combined agreement with a “C” grade: *Cost per test-positive participant* (*Health System*), corresponding to a 42.9% agreement.

The ranking of the indicators, shown in Table 3, reflects expert-identified priorities within each domain. The five highest ranked indicators per domain were: *“Screening acceptability”* (Prevention) *“Prevalence of HBV infection”* (Testing), *“Prevalence of HCV active infection”* (Morbidity), *“Onsite communication of rapid test results”* (Linkage to care), *“HCV Treatment initiation”* (Treatment & Care), *“Participation acceptability”* (Health system). A subset of the 18 indicators, i.e. the three highest-ranked per domain, is presented in Box 2 as a potential core set for practical application.

##### Box 2: List of core indicators

|  |
| --- |
| ***PREVENTION*** |
| Screening acceptability |
| HBV vaccination coverage (at least one dose) |
| HBV vaccination coverage (complete regime) |
| ***TESTING*** |
| Prevalence of HBV infection |
| Prevalence of current or past HCV infection (based on HCV-antibody testing) |
| HCV RNA/HBV DNA testing |
| ***MORBIDITY*** |
| Prevalence of active HCV infection |
| Prevalence of end-stage liver disease |
| Prevalence of liver cancer |
| ***LINKAGE TO CARE*** |
| Onsite communication of rapid test results |
| Referral of positive participants |
| Linkage to care among positive participants |
| ***TREATMENT AND CARE*** |
| HCV Treatment initiation |
| HCV treatment completion |
| HBV Treatment initiation |
| ***HEALTH SYSTEM*** |
| Sampling proportion |
| Participation acceptability |
| Self-reported barriers to access care |

## DISCUSSION

Using a two-round Delphi process, this study identified and prioritised a set of 50 key indicators for monitoring and evaluating screening, prevention, and management interventions for viral hepatitis B and C in community settings among migrants and refugees. The indicators were clustered into six domains *prevention, testing, morbidity, linkage to care, treatment and care*, and *health system*, with a consistently high level of expert agreement. These indicators respond directly to calls for “micro□elimination”, the strategy of achieving hepatitis elimination in clearly defined sub□populations through focused, data□driven action (46). Migrants and refugees are recognised micro□elimination target populations due to their often geographically clustered presence, accessibility through community networks, and underrepresentation in national surveillance systems (47). The indicators proposed through this Delphi process align with the core monitoring outcomes identified by Lazarus et al. (46) for HCV micro-elimination efforts, including prevalence of infection, people living with a diagnosis, treatment initiation and cure; because these interventions can show progress over a short time□frame, national or regional programmes can use them to demonstrate quick, measurable “small wins,” building confidence and momentum for wider hepatitis□elimination policies. In parallel, the 2024 Lancet Commission update identified simplified models of care, better community-based diagnostic strategies, and rigorous indicator frameworks as prerequisites for closing the persistent diagnosis and treatment gaps to eliminate viral hepatitis (1). By aligning with those priorities, our set can contribute to national programmes in quantifying how migrant□focused services impact overall elimination targets and facilitate comparisons across jurisdictions while guiding resource allocation.

The *Testing* and *Morbidity* domains recorded the highest proportion of unanimously accepted indicators, reflecting their objective measurability (48,49) and concordance with international guidelines (5,26). Conversely, the *Health System* domain showed the least consensus, suggesting the need for further exploration of proxies for the effectiveness of support services and the economic sustainability of community initiatives. In the *Prevention* domain, indicators related to self-reported testing and risk behaviours were rated less favourably, probably, most likely due to recall and social desirability biases which undermine their reliability (50).

The highest-ranked indicators include screening acceptability, prevalence of viral infections and liver cancer, communication of test results, referral to specialist centres and treatment initiation for HBV and HCV. These mark the critical steps of the diagnostic to care cascade. “*Screening acceptability”* reflects the willingness to undergo testing, the effectiveness of awareness campaigns, and the remaining economic, cultural, or logistical barriers (51,52). Early diagnosis and timely communication of results in HBV and HCV screening in community settings are essential to measure the disease burden and ensure rapid access to care, reducing the risk of loss to follow-up, and to available treatments, preventing progression (53–56). “*Prevalence of liver cancer”* reflects missed opportunities for early diagnosis and treatment, while also demonstrating screening’s capacity to detect advanced disease. Finally, referral to specialist care and treatment initiation assess how well health systems channel HBV□ or HCV□positive individuals into specialised management pathways that prevent disease progression and improve health outcomes (57).

However, some indicators showed some discrepancy between rating and ranking. For instance, “*HBV vaccination coverage (at least one dose)”* and *“HBV vaccination coverage (complete regimen)”* received a high ranking despite a moderate grade of agreement (respectively B, 85.7%, and A, 92.9%). This gap may be due to operational challenges, as vaccinations are often administered in health facilities (54), making coverage in the community more difficult to track and increasing dropout risk. Nonetheless, their high ranking confirms their strategic importance, regardless of implementation barriers. Similarly, “*Sampling proportion”*, scored low (B, 78.6%), yet was ranked as a priority, suggesting that participant recruitment would be considered a critical factor for the overall success of interventions. Nevertheless, denominators, expected numbers and invitation logs are often uncertain in community settings and may explain the lower agreement among experts despite its high perceived importance.

Indicators on continuity of care and treatment outcomes (e.g., “*HCV treatment success”*, “*Retention in care”, “Additional assessment among positive participants*)” achieved good agreement but lower rankings, possibly because they are very relevant but only emerge in long-term evaluations and may not entirely feasibile to monitor in community settings.

Research and policy documents have summarised and evaluated the hepatitis C control best practices (58), and international (5,26), and national (59,60) monitoring of viral hepatitis. Also, community interventions for high-risk groups have been assessed (61–63). However, no study has yet defined and validated a specific indicator set for monitoring community-based programmes aimed at migrants and refugees. These populations carry a disproportionate HBV/HCV burden and face legal, linguistic, cultural, and economic barriers to timely prevention and care. The lack of standardized monitoring tools perpetuates a fragmented and reactive (rather than anticipatory) response. This study addresses this gap by providing a structured framework for evaluating and optimizing interventions for these populations, contributing to a more effective, evidence-based response. Systematic impact measurement facilitates reporting and advocacy, equipping policymakers and stakeholders with concrete data for strategic decisions and inclusive policies. Standardized indicators enhance coordination among local, national, and international programmes, guiding resources to interventions which can be more cost-effective. Given that data should be collected with uniform definitions, this would yield accurate, comparable intelligence, highlighting gaps in screening or care and underpinning flexible, real□time planning that can adapt to epidemiological or operational shifts. By limiting the dashboard to key performance indicators that serve as proxies for success in each domain, evaluators can focus on the aspects that matter most while still obtaining a comprehensive picture of impact (44).

### Strengths and limitations

The major strength of this study lies in its novelty as the first effort to propose a standardized set of indicators for monitoring and evaluating viral hepatitis B and C screening, prevention, and management in community programs among migrants and refugees. The Delphi methodology employed ensures that the selected indicators are firmly grounded in scientific evidence while also incorporating expert consensus. However, consensus-building presents inherent challenges, particularly due to potential variability in expert perspectives and the need to balance diverse priorities within the selection process.

One key limitation concerns the expert selection process. Although convenience sampling may have introduced recruitment bias and limited diversity, we carefully assembled a panel with varied expertise to provide a well-rounded perspective on different aspects of viral hepatitis prevention. Furthermore, while response rates are often a concern in Delphi studies, a recent systematic review (64) noted that few studies report response rates across all rounds and indicated that the median number of invited experts is only 17 participants. In the present study, 41 experts were invited and 14 provided complete responses for both rounds. In light of these findings, the response rate can be considered acceptable, although not as high as initially anticipated. Nevertheless, the composition of the responding panel may have influenced the prioritization of indicators. Among those who completed the Delphi process, 65% were academic experts, which likely strengthened the scientific rigor of the evaluation but may have also placed greater emphasis on research-driven aspects over operational feasibility. Furthermore, since most respondents were based in Europe, adaptations may be necessary to ensure these indicators remain applicable in other global settings. Even so, the iterative Delphi process allowed experts to reassess and refine their evaluations across multiple rounds, helping to mitigate potential biases and enhance the robustness of the final selection. Indicators originated from a scoping review, so metrics absent from the published literature may have been overlooked. Nevertheless, the systematic approach offered a transparent evidence base, and open-ended questionnaire fields enabled experts to propose new items or revisions, helping bridge potential gaps.

### Future considerations

Future research should validate these indicators in operational programmes, paying particular attention to health□system and linkage□to□care metrics, where data availability and interoperability differ widely. Regional adaptations may also be necessary to reflect variations in health policy, access barriers and disease burden outside Europe and North America. Moreover, implementation studies should quantify the resources required to collect each indicator so that monitoring remains sustainable in low□resource settings.

## CONCLUSION

This consensus□based framework offers a practical, evidence□grounded tool for tracking the performance of community HBV/HCV interventions targeting migrant and refugee populations. While further validation is needed to assess their implementation across diverse contexts, these indicators provide a structured framework for improving data-driven decision-making and optimizing community-based strategies. Ultimately, their adoption could enhance the effectiveness of viral hepatitis screening, prevention, and management, contributing to broader public health efforts toward elimination goals.

## AUTHOR CONTRIBUTION

Conceptualisation: DP, AN, AT, CAP, MDP, VT, FDB, CLV, JAPM, GC, JVL, AMP, SB. Methodology: DP, AN, CAP, AMP. Project administration: DP, AN, AMP. Data collection, curation, and validation: DP, AN, VT, CAP, FDB. Formal analysis: DP, AN, CAP, AMP. Writing—original draft: DP, AN, AMP. Writing—review and editing: All authors; Supervision: CAP, AMP, SB. Final approval was obtained from all authors. All authors confirm that they had full access to all the data in the study and accept responsibility to submit for publication.

## DATA AVAILABILITY STATEMENT

All data produced in the present work are contained in the manuscript and supplementary materials.

## FUNDING

This study was co-financed through the Multi-country Viral Hepatitis COMmunity Screening, Vaccination, and Care project (VH-COMSAVAC). VH-COMSAVAC was co-funded by the European Union, under the EU4Health Programme (EU4H) (Project No. 101079958). Views and opinions expressed are those of the authors only and do not necessarily reflect those of the European Union or the European Health and Digital Executive Agency. Neither the European Union nor the granting authority can be held responsible for them.

## DECLARATION OF INTERESTS

JVL received grants to his institutions from AbbVie, Boehringer Ingelheim, Echosens, Gilead Sciences, Madrigal Pharmaceuticals, Moderna, MSD, Novo Nordisk, Pfizer, and Roche Diagnostics, consulting fees from Echosens, Global NASH Council, GSK, Madrigal Pharmaceuticals, Novo Nordisk, and Pfizer, and honoraria for lectures from AbbVie, Echosens, Gilead Sciences, GSK, Janssen, Moderna, MSD, Novo Nordisk, Pfizer, and Prosciento outside of the submitted work. CAP reports consulting fees from Roche Diagnostics and GSK, unrelated to this work. All other authors declare they have no competing interests.

## ACKNOWLEDGEMENTS

The authors would like to thank all experts who provided valuable input through the Delphi process of this study. AN, CAP and JVL acknowledge support to ISGlobal from the grant CEX2023-0001290-S funded by MCIN/AEI/ 10.13039/501100011033, and support from the Generalitat de Catalunya through the CERCA Programme.

## DECLARATION ON THE USE OF AI

AI tools (i.e., ChatGPT, Gemini) were used to improve the readability and language of the manuscript. The authors take full responsibility for the content and accuracy of the work.

