## Supplementary Materials for "Monitoring and evaluation of community interventions for viral hepatitis among migrants and refugees: a Delphi-based study"

* co-first authors

\# co-senior authors

1. **Amendement to the protocol**
2. **Systematic step-by-step list for the Delphi process**
3. [**Delphi studies in social and health sciences – recommendations for an interdisciplinary standardized reporting (DELPHISTAR) checklist** **(link to the Excel workbook)**](https://osf.io/nj435)
4. [**List of articles and initial list of indicators (link to the Excel workbook)**](https://osf.io/dyqp6)
5. [**List of indicators for Round 1 (link to the Excel workbook)**](https://osf.io/kj7n3)
6. [**Characteristics of panellists and results of Round 1 (link to the Excel workbook)**](https://osf.io/sncya)
7. [**List of indicators for Round 2 (link to the Excel workbook)**](https://osf.io/53z9n)
8. [**Results of Round 2 (link to the Excel workbook)**](https://osf.io/dkc75)
1. **Amendments to the protocol – November 27, 2024**

Monitoring and evaluation of viral hepatitis B and C screening, prevention and management: a protocol for prioritisation of performance indicators for community programmes targeting migrants and refugees.

Published on OSF on 5 November 2024, and accessible at <https://osf.io/m7jz3/>

All changes are listed below

| **Manuscript section** | **Amendment/Clarification** | **Rationale** |
| --- | --- | --- |
| Data extraction | Indicators will be identified by thoroughly reviewing all sections of each article, including the abstract, methods, results, and discussion. Specific attention will be given to any explicitly reported indicators, as well as data points that could be used to derive relevant measures. Furthermore, tables, figures, and supplementary materials will be examined to ensure a comprehensive extraction of indicators. | The amendment clarifies the indicator extraction process by specifying a comprehensive review of all article sections, including tables and supplementary materials. |
| Prioritization of indicators | To enhance the rigor and comparability of this Delphi study, the DELPHISTAR reporting guideline will be applied, following its recommendations for structured reporting in health and social sciences research | The amendment includes a reference to the DELPHISTAR reporting guideline to ensure standardized and transparent reporting of the Delphi process. |
| Delphi method data collection | Indicators that achieved 67% or less agreement for responses of "agree" or "somewhat agree" (combined agreement) in R1 will be excluded from further consideration. Conversely, those exceeding this threshold will proceed to R2. Indicators that do not undergo substantial modifications will advance directly to the ranking phase, while those that undergo significant revisions will be re-evaluated through an additional rating process before moving forward.  During R1, panellists will be required to assess whether the indicators from the additional list—representing those for which the core group has not reached a consensus—should be incorporated into the main list. This evaluation will be conducted using a simple yes/no response format. Indicators receiving at least 67% affirmative votes will be included in the main list and rated further in R2.  In R2, panellists will review the revised indicators along with summaries outlining the modifications made. Indicators that undergo substantial changes following R1 feedback, as well as those selected from the additional list, will be subjected to re-evaluation. Using the same four-point Likert scale as in R1, panellists will reassess these indicators to determine their suitability for inclusion in the final list. Separately, panellists will rank the indicators within each health domain based on their practical applicability in community-based programs for viral hepatitis B and C screening, prevention, and management among migrants and refugees.  For the ranking, panellists will arrange the indicators in order of relevance, from the most to the least important.. If an indicator is considered not relevant, they will have the option to choose ‘I prefer not to rank this indicator.’ Experts will be given the opportunity to suggest minor edits to enhance the clarity of the indicator without changing its meaning, while feedback requiring substantial modifications will not be taken into account. | The amendment provides a clearer specification of the rating and ranking process, detailing how indicators progress through different evaluation phases and how panellists assess their relevance and applicability. |
| Delphi data analysis | The quantitative analysis of R1 and R2 results will focus on the rating of each indicator, based on responses provided by panelists using a 4-point Likert scale. Each panelist will rate the indicator across the five predefined items (Box.1), assigning scores from 1 (‘Disagree’) to 4 (‘Agree’), with an additional category for 'Not qualified to respond'.  To obtain a single summary measure reflecting each participant’s overall assessment of the five evaluation criteria, a total score for each expert will be calculated for each indicator. Each indicator will be rated across five items (Box 1) using a 4-point Likert scale (1 = “Disagree”, 4 = “Agree”), and the scores assigned to the five items will be summed to generate an individual total score (for each expert). This score will range from a minimum of 5 (all responses “Disagree”) to a maximum of 20 (all responses “Agree”).  To classify the final overall level of agreement, the total score of each expert (for each indicator) will be mapped onto one of four predefined agreement categories (“Agree”, “Somewhat agree”, “Somewhat disagree”, “Disagree”). This classification will be carried out by calculating the score range and dividing the total interval (5–20) into four equal segments of 3.75 points, each corresponding to a specific level of agreement.  A detailed example of the scoring system is provided below:   - The total score will range from 5 (all responses “Disagree”) to 20 (all responses “Agree”) - Range = 20 – 5 = 15 - Each agreement category will correspond to an equal range of 15 / 4 = 3.75 points   Agreement categories and score thresholds:   - Agree: 16.25 ≤ Score ≤ 20.00 - Somewhat agree: 12.50 ≤ Score < 16.25 - Somewhat disagree: 8.75 ≤ Score < 12.50 - Disagree: 5.00 ≤ Score < 8.75   If a participant selects the “Not qualified to respond” option for one or more items, those responses will be excluded from the total score calculation. In such cases, the maximum possible score range will be proportionally adjusted according to the number of valid responses, to ensure consistency in the interpretation of agreement levels across indicators with different response counts.  In addition, once the individual total scores are obtained, the percentage of responses falling into the categories “Agree”, “Somewhat agree”, “Somewhat disagree”, “Disagree”, or “Not qualified to respond” will be calculated for each indicator to assess the overall level of agreement. | The amendment provides further details on the calculation of rating and ranking, specifying how standardized indicator-level scores are derived, how agreement levels are categorized, and how ranking is determined based on mean scores. |

**Systematic step-by-step list for the Delphi process**

**To enhance clarity and readability, the key steps undertaken during each phase of the Delphi process are systematically presented below.**

### ***Round 1 (R1)***

- Panellists rated the preliminary main list of indicators using a four-point Likert scale.
- Indicators that received **more than 67% combined agreement** (“agree” or “somewhat agree”) proceeded to R2, while those with **67% or less** were excluded from further consideration.
- Panellists were also asked to evaluate a **separate set of indicators**—the additional list—which included indicators for which the core research team had not reached consensus during the preparatory phase.
  - These additional indicators were assessed using a **yes/no** format to determine whether they should be included in the main list.
- Indicators from the additional list that received **at least 67% affirmative votes** from the panel were added to the main list and included in R2.Sociodemographic and professional characteristics of panellists (e.g., gender, age, country, job category, and level of expertise) were systematically collected during R1.Panellists also had the opportunity to provide open-ended feedback to support indicator refinement, if deemed necessary.

***Between R1 and R2***

- We reviewed expert feedback and revised indicators accordingly.
- **Indicators with only minor edits** (e.g. wording or clarity) were **not re-rated** but moved directly to the ranking phase.
- **Indicators that underwent substantial revisions** (e.g. changes in meaning or structure) were flagged for **re-evaluation,** through an additional rating process**,** in R2.

### ***Round 2 (R2)***

- Panellists reviewed summaries of the modifications made. The feedback was aggregated across all expert groups.
- Indicators that had undergone **substantial changes** after R1, along with those newly added from the additional list, were **re-evaluated** using the same four-point Likert scale.
- **Separately**, panellists were asked to **rank the indicators** within each health domain based on their **practical applicability** in community-based HBV/HCV programmes for migrants and refugees.
  - Panellists arranged the indicators **in order of relevance, from the most to the least important**.
  - If an indicator was considered not relevant, they could select the option **“I prefer not to rank this indicator.”**
- Experts were allowed to suggest **minor wording edits** to improve clarity, while feedback requiring major revisions was not considered at this stage.
